# Epidemiological Analysis of Lower Limb Revascularization for Peripheral Arterial Disease over 12 years in the Public Health System in Brazil

**DOI:** 10.1101/2021.11.05.21265468

**Authors:** Nelson Wolosker, Marcelo Fiorelli Alexandrino da Silva, Maria Fernanda Cassino Portugal, Nickolas Stabellini, Antônio Eduardo Zerati, Claudia Szlejf, Edson Amaro, Marcelo Passos Teivelis

## Abstract

**Objectives:** Worldwide, peripheral arterial disease (PAD) is a disorder with high morbidity, affecting more than 200 million people. Our objective was to analyse the surgical treatment for PAD performed in the Unified Health System of Brazil over 12 years based on publicly available data.

**Methods:** The study was conducted with data analysis available on the DATASUS platform of the Brazilian Health Ministry, assessing procedure technique distribution throughout the years, mortality and cost.

**Results:** A total of 129,424 procedures were analysed (either for claudicants or critical ischemia, proportion unknown). The vast majority of procedures were Endovascular (65.49%), with a tendency for increase in this disproportion (p<0.001). There were 3,306 in-hospital deaths (mortality of 2.55%), with lower mortality in the Endovascular group (1.2% vs. 5.0%, p=0.008). The overall governmental investment for these procedures was U$ 238,010,096.51, and Endovascular Procedures were on average significantly more expensive than Open Surgery (U$ 1,932.27 v. U$ 1,517.32; p=0.016).

**Conclusions:** In the Brazilian Public Health System, lower limb revascularizations occurred with gradual growing frequency between 2008 and 2019. Endovascular procedures were vastly more common, and related to lower in-hospital mortality rates, but higher procedural costs.

## Introduction

Estimations assume, should current trends remain, that over 20 million deaths will occur worldwide from cardiovascular diseases (CVD) by 2030(1). The more remarkable burden is anticipated for low and middle-income countries (LMICs), mostly due to the expected population growth(2,3).

Peripheral Arterial Disease (PAD), often marked by an altered ankle-brachial index(4,5), is a well-established predictor of cardiovascular mortality(6), estimated to affect over 200 million people worldwide(7), and to which over 40,000 yearly deaths were attributable in accordance with the 2013 Global Burden of Disease Study, representing an 155% increase from the year 1990(2). Although diagnosis is simple and the management approachable and feasible, PAD remains relatively under-recognized and undertreated(6). Management should always include risk control with lifestyle changes and medical therapy, also currently comprising surgical or endovascular options of revascularization, with a marked increase in the use of endovascular techniques in the past two decades(8).

A comprehensive study by Gheorghe and collaborators, in which authors aimed to determine the knowledge gaps to the economic burden of CVD in LMICs, identified a heterogeneous body of literature, dominated by single centre retrospective cost studies conducted in secondary care settings(9). Specifically in Brazil, the authors report finding only 8 papers which offer economic estimates with regard to CVD(9). With regard to PAD, a Brazilian study published in 2016 has shown a high economic impact and a growth trend for the endovascular technique, with a 57% increase in endovascular procedures between 2008 and 2012(10). One single other study, published by Wolosker *et al*, performed a descriptive analysis of the treatment of PAD in the largest Brazilian city over the spam of 12 years, encompassing 10,951 procedures(11). These authors found that, in the city of São Paulo, endovascular procedures were more common than open surgeries and resulted in shorter hospital stays as well as lower perioperative mortality rates(11). No study, however, has been conducted which analysed the management of PAD in the whole Brazilian territory.

Brazil is a country whose estimated population for the year 2020 is 211,755,692 inhabitants(12). Of these, over 75% were, as of June 2020, solely dependent on the Public Health System (SUS), which is a tax-funded governmental service; whereas 24.1% benefited from private supplementary health care, individually financed or maintained by an employer(13). In this scenario, this study aims to present the first descriptive analysis of the management of PAD in the entire Brazilian territory, ranging from 2008 to 2019, assessing procedure technique distribution throughout the years, mortality and cost.

## Methods

All information was extracted from the DATASUS portal, which is a digital platform though which the Brazilian government provides open data relating to procedures performed under the Public Health System in adequately accredited hospitals. Institutional accreditation in the system is prerequisite for receiving governmental payment for the procedures performed. All data is appropriately de-identified.

This study was submitted to our institutional review board at the Hospital Israelita Albert Einstein (protocol number 4324-20). Since all data was anonymous, a waiver of Informed Consent was requested and granted.

Study was approved by the Hospital Israelita Albert Einstein Ethics Committee under protocol 35826320.2.0000.0071 by Technical Statement 4.321.508, issued on the 6^th^ of October, 2020.

Data referring to open surgical and endovascular procedures for treatment of PAD between 2008 and 2019 was selected from the platform, along with information regarding procedure technique, mortality and cost. The reported values refer to the costs of hospitalization and include the surgical devices.

A total of 14 procedure codes were elected for procedure listing, selected from the SUS Table of Procedures, Medicines and OPM Table Management System (SIGTAP): Open Surgery procedures: 04.06.02.031-0 - Axillobifemoral bypass; 04.06.02.032-9 - Axillofemoral bypass; 04.06.02.034-5 - Crossed femoro-femoral bypass; 04.06.02.035-3 - Aorto-femoral thromboendarterectomy; 04.06.02.036-1 - Aorto-iliac bypass/thromboendarterectomy; 04.06.02.038-8 - Iliac-femoral bypass/thromboendarterectomy; 04.06.02.043-4 - Distal artery revascularization bypass or thromboendarterectomy; 04.06.02.044-2 - Distal artery revascularization bypass or thromboendarterectomy from the proximal femoro-popliteal segment; 04.06.02.045 - Femoro-popliteal proximal bypass Revascularization or thromboendarterectomy. Endovascular procedures: 04.06.04.028-1 - Aortic intraluminal angioplasty, vena cava/iliac vessels (with stenting); 04.06.04.004-4 - Intraluminal aortic angioplasty, vena cava/iliac vessels (without stent); 04.06.04.005-2 - Intraluminal vessel angioplasty of distal artery (without stent); 04.06.04.006 - Intraluminal vessel angioplasty of distal artery (with uncovered stent); 04.06.04.002-8 - Reconstruction of the aortoiliac bifurcation with angioplasty and stenting.

Revascularization procedures were divided into two groups: Open Surgery and Endovascular Procedures. As is common with population code-based studies, miscoding or data loss may have occurred; considering the bulk of the sample, however, this is not expected to negatively impact the overall results.

All data was collected through an automated web scraping method. The codes employed were programmed in Python language (v. 2.7.13; Beaverton, OR, USA), using the Windows 10 Single Language operating system. Field selection in the DATASUS platform and posterior table adjustment were performed by Selenium WebDriver packages (v. 3.1.8; Selenium HQ, various contributors worldwide) and pandas (v. 2.7.13; Lambda Foundry, Inc. and PyData Development Team, NY, USA). All data was organized into Microsoft Office Excel 2016® (v. 16.0.4456.1003; Redmond, WA, USA) spread sheets after collection and treatment.

The monetary values in Brazilian Reais were converted into U.S. dollars using the exchange rate of the 31st of December, 2012, which is the median date of the cases evaluated, and corresponds to U$ 1 = R$ 2,04.

Statistical analysis was as follows: the Chi-square test was used for determination of a trend in distribution of procedure techniques throughout the years, and groups were compared with regard to mortality rates and average costs by the Mann-Whitney test. For all tests, the level of statistical significance was < 0.05.

## Results

In total, 129,424 procedures for the revascularization of PAD were performed in Brazil between 2008 and 2019, with a yearly average of 10,785.33 procedures.

Considering this number and the approximate 160.722.570 inhabitants dependent on the Public Health System in Brazil in 2020, the procedure rate was 6.7 procedures per 100,000 inhabitants per year.

The frequency of procedure technique throughout the years is shown in Table 1. The majority of procedures were done by Endovascular technique, with a technological transition observed especially from 2010 on, with a significant tendency to increase the number of endovascular procedures to the detriment of Open Surgery (*p*<0.001).

**Table 1:** Absolute and relative frequency of endovascular and open procedures for DAP between 2008 and 2019

|  | Open Surgery |  | Endovascular Surgery |  | Total | *p* * |
| --- | --- | --- | --- | --- | --- | --- |
|  | n | % | n | % |  |  |
| 2008 | 4,595 | 56.33 | 3,562 | 43.67 | 8,157 | <0.001 |
| 2009 | 4,370 | 51.75 | 4,075 | 48.25 | 8,445 |  |
| 2010 | 4,233 | 49.58 | 4,305 | 50.42 | 8,538 |  |
| 2011 | 4,088 | 44.99 | 4,998 | 55.01 | 9,086 |  |
| 2012 | 3,913 | 41.81 | 5,446 | 58.19 | 9,359 |  |
| 2013 | 3,765 | 35.57 | 6,821 | 64.43 | 10,586 |  |
| 2014 | 3,483 | 30.17 | 8,061 | 69.83 | 11,544 |  |
| 2015 | 3,325 | 27.21 | 8,897 | 72.79 | 12,222 |  |
| 2016 | 3,440 | 27.17 | 9,219 | 72.83 | 12,659 |  |
| 2017 | 3,374 | 25.91 | 9,649 | 74.09 | 13,023 |  |
| 2018 | 3,297 | 24.13 | 10,369 | 75.87 | 13,666 |  |
| 2019 | 2,781 | 22.91 | 9,358 | 77.09 | 12,139 |  |
| **Total** | **44,664** | **34.51** | **84,760** | **65.49** | **129,424** |  |

\* trend chi-square test

Mortality data is depicted on Table 2. There were 3,306 in-hospital deaths (mortality rate of 2.55). The in-hospital mortality rate for Endovascular procedures was lower than that for Open Surgery (1.2% *v*. 5.0%, *p*=0.008).

**Table 2:** Absolute and relative mortality per geographic region by procedure type

| Geographic Region | Total |  |  | Procedure Type |  |  |  |  |  |
| --- | --- | --- | --- | --- | --- | --- | --- | --- | --- |
|  | Procedures | Mortality |  | Open Surgery |  |  | Endovascular Surgery |  |  |
|  |  | n | (%) | Procedures | Mortality n | (%) | Procedures | Mortality n | (%) |
| North | 1,978 | 62 | 3.13 | 560 | 34 | 6.07 | 1,418 | 28 | 1.97 |
| Northeast | 19,799 | 447 | 2.26 | 4,455 | 256 | 5.75 | 15,344 | 191 | 1.24 |
| Southeast | 56,821 | 1,650 | 2.90 | 22,446 | 1239 | 5.52 | 34,375 | 411 | 1.20 |
| South | 45,689 | 976 | 2.14 | 15,032 | 624 | 4.15 | 30,657 | 352 | 1.15 |
| Center-West | 5,137 | 171 | 3.33 | 2,171 | 104 | 4.79 | 2,966 | 67 | 2.26 |
| **Total** | **129,424** | **3,306** | **2.55** | **44,664** | **2,257** | **5.05** | **84,760** | **1,049** | **1.238** |
| *p* * |  |  |  |  |  |  |  |  | 0.008 |

\* Mortality in the Open Surgery group was statistically higher than in the Endovascular Surgery group (p = 0.008), Mann-Whitney test.

Costs covered by the Public Health System are presented in Table 3. The total expense for all procedure throughout the evaluated years was U$ 238,010,096.51. Endovascular Procedures were significantly more expensive than Open Surgery (27.3% more expensive, U$ 1,932.27 *v*. U$ 1,517.32; *p*=0.016).

**Table 3:**
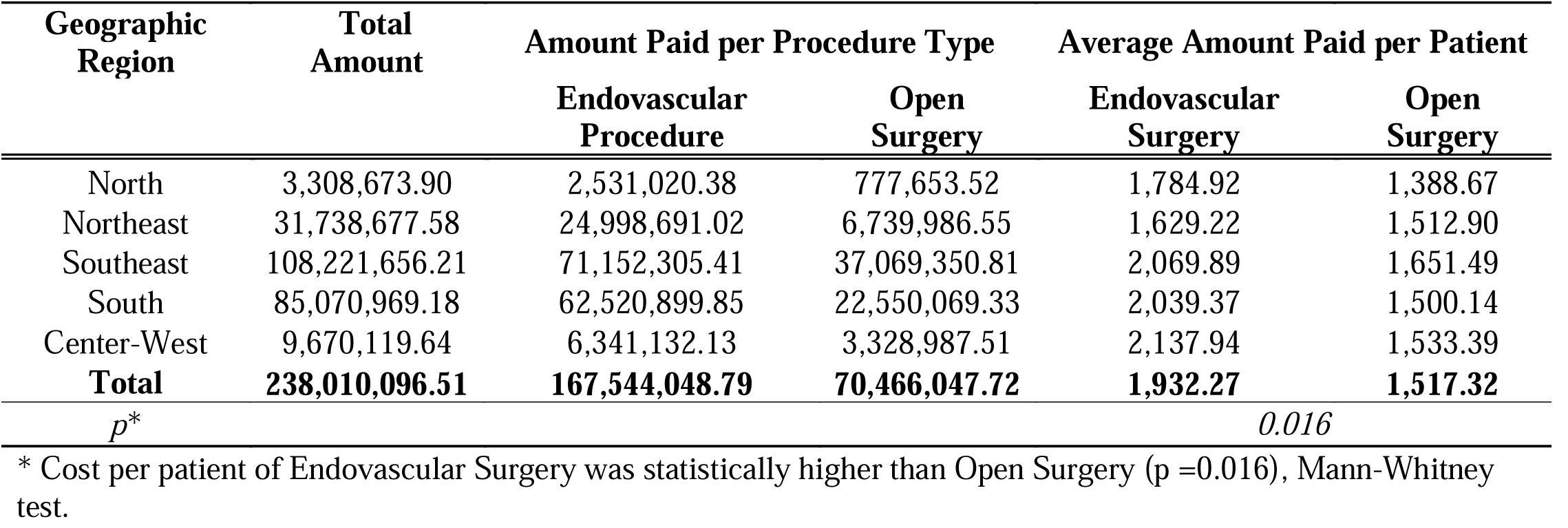
values paid by SUS in US dollars per geographic region by procedure type

| Geographic Region | Total Amount | Amount Paid per Procedure Type |  | Average Amount Paid per Patient |  |
| --- | --- | --- | --- | --- | --- |
|  |  | Endovascular Procedure | Open Surgery | Endovascular Surgery | Open Surgery |
| North | 3,308,673.90 | 2,531,020.38 | 777,653.52 | 1,784.92 | 1,388.67 |
| Northeast | 31,738,677.58 | 24,998,691.02 | 6,739,986.55 | 1,629.22 | 1,512.90 |
| Southeast | 108,221,656.21 | 71,152,305.41 | 37,069,350.81 | 2,069.89 | 1,651.49 |
| South | 85,070,969.18 | 62,520,899.85 | 22,550,069.33 | 2,039.37 | 1,500.14 |
| Center-West | 9,670,119.64 | 6,341,132.13 | 3,328,987.51 | 2,137.94 | 1,533.39 |
| **Total** | **238,010,096.51** | **167,544,048.79** | **70,466,047.72** | **1,932.27** | **1,517.32** |
| *p\** |  |  |  | *0.016* |  |

\* Cost per patient of Endovascular Surgery was statistically higher than Open Surgery ( $p=0.016$ ), Mann-Whitney test.

## Discussion

All data in this study was obtained from publicly available websites by use of web scraping codes. Manual collection of this volume of data would be rather industrious, though technically feasible. Automated collection, on the other hand, permits acquiring information in a faster and uncomplicated fashion. It must be noted, however, that this process is not exempt from coding errors, especially considering the fact that all information is sourced from an administrative databank instead of medical records, as well as the fact that the coding system may permit a certain ambiguity of codes for procedures employed for different diseases, such as revascularizations for PAD and trauma. Since the diagnosis cannot be tracked from the de-identified database, exclusion of unwanted cases was not possible; but it may be surmised that, because of the bulk of cases, a very small part of the information is wrongly considered. Unfortunately, unlike the municipal platforms, the nationwide platform does not permit exclusion of disease codes, which precludes the omission of trauma cases (International Classification of Diseases 10th revision – ICD-10 – S or T) submitted to revascularization. In order to solve this issue, the proportion of cases subjected to revascularization under ICD-10 diagnostics S or T – that is, patients treated for trauma, instead of PAD – was evaluated for the city of São Paulo, which is Brazil’s largest urban centre, based on the casuistry of Wolosker(11). In São Paulo, between 2008 and 2018, only 0.14% of all revascularizations were classified as being a consequence of trauma. Thus, if this proportion may be applied to the entirety of Brazil’s territory by analogy, it could be assumed that the number of non-excludable trauma cases in this sample is of little relevance.

Our data demonstrates a gradual increase in the number of patients treated throughout the evaluated years, with an average of 10,785.33 procedures per year. Information regarding the incidence of PAD worldwide is relatively lacking(14). A yearly average of 6,283.25 open revascularizations was found in England from data collected between 2002 and 2006 from the Hospital Episode Statistics and Office for National Statistics in a study by Moxey and collaborators(15). Considering an estimated population of 50,97 million people in England in 2006(16), the rate of open procedures was 12.32 per 100,000 inhabitants, in contrast with 27.79 open procedures per 100,000 inhabitants in Brazil. In the UK, however, the incidence of PAD decreased steadily between 2000 and 2014, from 38.55 to 17.33 per 10,000 person-years(17), whereas results of a recent meta-analysis indicated that the prevalence of PAD increased from 2000 to 2010 in high-income countries(14). Long-term results from the German ‘getABI’ study, which enrolled patients aged 65 years and older, determined a PAD incidence of 203 per 10,000 person-years(18).

In a study of French patients hospitalized for PAD under the National Health Insurance Scheme between 2007 and 2011, revascularizations were performed in 9.5% of cases, from a total of 7,266 evaluated patients(19). In our study, a high volume of treated patients and an increasing number of procedures per year points to either an increase in disease incidence or an increment in governmental investments for the management of PAD.

There was, in our sample, a clear predominance of endovascular procedures (65.49%), which occurs all through the studied years, except 2008 and 2009 (43.67 and 48.25%, respectively), indicating a technological transition around the year 2010. In the study by Wolosker *et al,* which focused only in the city of São Paulo, this technological transition with an inversion between the frequencies of open surgery and endovascular procedures seems to have happened earlier, even before 2008(11). In a study performed in a teaching hospital in São Paulo, the technological transition from endovascular procedures to open surgery was found beginning as early as 2006(20).

This expansion in the application of endovascular technique is in accordance with observations by other authors(21), and may be attributed to several reasons: the most conspicuous is, naturally, the relatively lower morbidity of endovascular procedures, which may broaden the spectre of treatable patients. Other authors suggest that a decrease in the threshold for intervention, with surgeries being performed for claudicant patients as well as those with critical limb ischemia, also represents a significant contribution to the increase in utilization of the endovascular technique(22,23). Some also propose that a rising awareness of the disease may be a contributing factor in the rising of the application of endovascular revascularization procedures(24). Finally, the fact that endovascular procedures may be less durable, thus requiring a number of reinterventions, may be a confounding factor when considering the frequency of techniques, especially because our de-identified dataset does not permit the exclusion of readmissions, thus limiting this evaluation. The databank anonymity also impairs determination of surgical indication, whether for claudication or critical limb ischemia.

With regard to in-hospital mortality, it was higher for Open Surgery than Endovascular Procedures (5.0% *v*. 1.2%, *p*=0.008). These findings are in accordance with a recent meta-analysis of twenty-seven trials (seven randomized controlled trials and twenty retrospective trials), encompassing 17,536 patients, conducted by Tang *et al*, which found higher mortality during follow up (10.86% *v*. 7.54%, p<0.05) in the open surgery group(25). The Bypass versus Angioplasty in Severe Ischemia of the Leg (BASIL) trial, which randomized 452 patients with severe limb ischemia due to infrainguinal disease to receive either a surgery-first or an angioplasty-first strategy, determined that at one-year open surgery and angioplasty did not differ significantly in amputation-free survival (71% bypass *v*. 68% angioplasty; adjusted HR = 0.73, 95% CI [0.49–1.07])(26), but at 5-years follow up bypass surgery was associated with better overall survival (47% bypass *v*. 41% angioplasty; adjusted HR = 0.61; 95% CI [0.50– 0.75]; P < 0.009) and a non-significant difference in amputation free survival (38% bypass *v*. 37% angioplasty; adjusted HR = 0.85; 95% CI [0.5–1.07]; P = 0.108)(27).

The mortality analysis in our sample, however, is limited due to two main factors: the fact that the procedure indication cannot be established from the available information, so patients operated on for claudication cannot be separated from those with critical limb ischemia, which may have an impact on mortality rates; and the fact that the databank information anonymization hinders patient follow-up impossible, with all mortality data discussed referring exclusively to in-hospital deaths; therefore, unable to contribute information with respect to long-term mortality rates.

In our sample, governmental cost for Endovascular procedures was higher than that for Open Surgery (U$ 1,932.27 *v*. U$ 1,517.32; *p*=0.016). Studies focusing on the cost of gross hospitalization, found that endovascular procedures in general represent shorter hospital stays and lower overall cost(26,28). The long-term economic assessment of the BASIL trial, however, showed that costs for Open Surgery were indeed higher in the first year ($34,378 for bypass *v*. $25,909 for angioplasty), but that at the end of a 5-year follow-up this difference decreased ($45,322 for bypass *v*. $39,801 for angioplasty) and was no longer significant, which authors associate with to costs subsequently incurred by the need of multiple reinterventions, including open surgery(26). It is important to remark, however, that the BASIL trial compared open surgery to plain angioplasty, and bulk of literature is lacking which refers to the cost-analysis of newer endovascular options including atherectomy and drug eluting devices(29). Another important assertion with regard to this analysis in Brazil, and possibly other LMICs, is that in countries where most endovascular devices are not locally sourced, additional costs such as import taxes and duties may apply(30).

## Limitations

As is pertaining to retrospective analyses, our study is limited by loss of patient information and eventual miscoding of an administrative database. Because only hospitals duly accredited in the SIGTAP registry feed the databank, there is certain to have been loss of information from those centres without accreditation which may have performed procedures in an urgency scenario.

Lastly, we were unable to provide input with regard to the need for reinterventions and the weight they bear in the frequency of endovascular procedures.

In spite of its’ limitations, this is a comprehensive analysis of the public health system’s management of PAD in one of the world’s largest and most populous LMICs, encompassing a 12-year interval and over 120,000 procedures, granting a representational assessment of a real-world sample of PAD patients. This study also presents a useful tool for better understanding of a public system and guidance for the allocation of health funds.

## Conclusions

The number of patients treated for PAD with surgical management by the Brazilian Public Health System increased gradually in the period between 2008 and 2019. Endovascular procedures were vastly more common, especially after a technological transition occurring in 2010. Endovascular procedures were associated with lower in-hospital mortality rates, at the expense of higher procedural costs (27.3% higher).

## Data Availability

All data produced in the present study are available upon reasonable request to the authors

